# Influences on rural HPV vaccine hesitancy: A multi-level mixed methods study

**DOI:** 10.1101/2024.12.29.24319671

**Authors:** Sherri Sheinfeld Gorin, Arturas Klugas, Matthew Stack, Samantha Chuisano, Hannah Knoll, Cornelius D. Jamison, Angela McGrady, Dongru Chen, Rebecca Hyman, Melissa DeJonckheere

**Affiliations:** University of Michigan Medical School University of Michigan School of Public Health 1018 Fuller St Ann Arbor, MI 48104, USA; DipABLM, FAAFP MSU/MyMichigan Medical Center Alma 300 Warwick Drive Alma, MI 48801, USA; MSU/MyMichigan Medical Center 300 Warwick Drive Alma, MI 48801, USA; University of Michigan Medical School 1018 Fuller St Ann Arbor, MI 48104, USA; Albert Einstein School of Medicine 1300 Morris Park Ave Bronx, NY, USA 10461

**Keywords:** HPV vaccine hesitancy, rural youth, multilevel, rural primary care physicians

## Abstract

Human Papillomavirus (HPV) is the most common sexually transmitted infection and the leading cause of cervical and oropharyngeal cancers. Vaccination can prevent over 90% of HPV- attributed cancers among those aged 9-26. Rural populations are less likely to receive the complete HPV protocol than urban. The purpose of this convergent mixed methods study was to explore in depth the perspectives that influence hesitancy for HPV vaccination among rural youth, caregivers, and primary care providers in rural Michigan. A mixed methods analysis was conducted to integrate quantitative (cross-sectional PCP survey) and qualitative data (youth, parent focus groups) in a joint display table. Youths and parents reported limited unbiased HPV vaccine information; yet, all responding physicians reported educating their patients about the HPV vaccine. Inductive descriptive analyses revealed that youth wanted a greater role in decision-making about the HPV vaccine, while parents were the PCPs primary focus.

Integrative analyses of both qualitative and quantitative data identified parental out-of-pocket costs as significant predictors of lower HPV vaccine uptake. Clinic structural supports were significantly associated with higher HPV vaccine uptake. Adapting evidence-based dissemination approaches to rural primary care physicians could decrease HPV vaccine hesitancy. Youths could be engaged as HPV vaccine champions. Accessing community resources could increase access.

## Introduction

In the United States, about 13,820 new cases of invasive cervical cancer will be diagnosed in 2024; about 4,360 women are estimated to die from this disease [1]. These relatively low mortality rates are attributable to the success of cervical cancer screening and the treatment of cervical intraepithelial neoplasia (CIN), earlier in the pathogenesis pathway to invasive cervical cancer [2–7]. Almost all cervical cancers are caused by persistent infections with oncogenic, or high-risk, types of human papillomavirus (HPV) [8]. Due to the rise in human papillomavirus (HPV)-associated oropharyngeal (oral cavity and pharynx) cancer, this cancer is increasing in incidence; men are more than twice as likely as women to be diagnosed [6, 9–18].

While HPV vaccines are effective against many of the oncogenic HPV types, vaccination uptake and completion are low among rural adolescents. The Advisory Committee on Immunization Practices (ACIP) has recommended routine administration of the HPV vaccine for 11-12 year olds, as early as age 9; catch up vaccines through age 26 [13–17, 19]. The up-to-date HPV vaccination rate in rural Michigan is lower than both the national and Michigan rates, at 26.09% [19], and has increased since 2013. Further, there is considerable vaccination variability within these rural midwestern counties[19, 20].

Vaccine hesitancy is defined by the World Health Organization as “delay in acceptance or refusal of vaccination despite availability of vaccination services.[21] Those who live in rural communities face unique challenges to HPV vaccination. Travel time and low density of healthcare resources may pose challenges for accessing primary care to administer the vaccine [22, 23]. Some rural public health programs do not have resources to promote HPV vaccination or to devote to immunization in general [24]. These resources---as well as HPV vaccination messages—may not be well-coordinated [23, 25–28]. Lack of awareness, limited knowledge, fear, and community stigma may increase hesitancy among adolescents and their caregivers [29–32]. These influences may vary across and within rural communities [33, 34]. for example, due to the diversity of residents, including in Michigan, among migrant agricultural workers [35, 36].

Multi-level approaches--with individuals, parents, clinical teams, health care institutions, engaged community stakeholders and policy-makers-- have been found effective in increasing HPV vaccine uptake [37]. For example, work by Walling, et al. [38], suggests that parental-level interventions may be effective in increasing HPV vaccine initiation. Numerous papers have found that primary care providers, particularly physicians, are key to HPV vaccine uptake, both initiation among rural populations [39] and series completion [39–46]. Recent studies have found that physicians’ attitudes and beliefs about HPV vaccine efficacy and safety may influence the rates at which they recommend vaccination to their patients [47].

At the community, level, a HPV Vaccination Van Program increased vaccine uptake in the rural communities in South Carolina. A recent study demonstrated the feasibility of enrolling a community pharmacy in a rural, medically underserved Alabama county as a Vaccines for Children (VFC) provider to provide free vaccines to eligible adolescents [48]. Recent studies demonstrate a positive impact of school mandate requirements—although relatively few nation- wide-- to the HPV vaccine [49–51]. Thus, multi-level interventions, addressing barriers at the patient, provider, community, and policy levels, could effectively increase HPV vaccine uptake[52] in rural communities.

The purpose of this convergent mixed methods study was to explore in depth the perspectives that influence hesitancy for HPV vaccination among rural youth, caregivers, and primary care providers in rural Michigan. Rural communities are heterogeneous, and studies of the multi-level influences on HPV vaccine uptake in these contexts are few [53].

## Materials and Methods

We conducted a convergent mixed methods study,[54] integrating qualitative findings from a series of focus groups held with children and adolescents, young adults and parents of participating children and adolescents with the results of a rural primary care physician survey. We sought to describe the perspectives of participants at multiple levels, and to identify complementary results from qualitative and quantitative data collection and analysis strategies.

### Setting

About 18% of Michigan residents live in rural areas [55]. Rural Michigan is poorer than urban Michigan; somewhat fewer individuals have completed high school; the unemployment rate in rural Michigan is the same as that for urban areas of the state.

Rurality has been defined variously by the OMB, the census bureau, and the USDA, among others. In this study, we applied the US census bureau definition of rurality [56, 57].

### Quantitative Methods

We administered an online survey about HPV vaccination practices, provider and parent attitudes and beliefs about HPV vaccine hesitancy to licensed rural primary care physicians in the State of Michigan [56]; there is no list of rural physicians using the US Census Bureau definition, however. Physicians who met the eligibility criteria as more than 50% time in clinical practice, and board certified in primary care (internal medicine, family medicine, pediatrics, general medicine) were sent a survey (N=49); they were also required to complete a two-page consent form via REDCAP. After targeted outreach to large rural practices, identified champions of rural health, and 5 follow-ups by email, 17 responses were received (35%). A low rate of response to survey (and other) research among primary care providers has been found in other studies post-COVID-19, even those that include additional resources (such as practice facilitation)[57], due to physician burnout. Rural primary care physicians are particularly difficult to recruit.[60] Among rural primary care physicians, the numbers of clinics have been reduced by 30% nationwide, further delimiting the pool of respondents.

The 32-item survey was administered by email, using REDcap. The primary outcome was vaccine hesitancy, measured for PCP’s as the continuous stages of change for inoculating age-eligible patients against the HPV; it ranges from 1, no thoughts of vaccine use and recommendation in my practice to 9, vaccinating and educating patients about HPV in my practice. The stages of change scale[61–65] has been widely used to measure behavior change, including in HPV vaccination, and has strong psychometric properties. The primary predictors included; provider attitudes/beliefs toward the HPV vaccine, clinician communication approaches, practice workflow for the HPV vaccination, parental attitudes/beliefs toward the HPV vaccine, and children’s and adolescents’ attitudes and beliefs toward the HPV vaccine. All items were derived from existing scales.

We conducted cognitive interviews with three practicing primary care physicians in rural Michigan during the pilot phase of the survey. The questionnaire was reduced in size, questions and scales were further refined.

### Qualitative Methods

#### Participants and Recruitment

Qualitative research seeks to gather data from information rich cases—those with lived experience relevant to the research question—to explore the shared perspectives that influence HPV vaccine hesitancy. By gathering information rich cases, the research team is seeking transferability rather than generalizability.[66] *Per force*, the goal of this sampling strategy was to look for shared experiences across stakeholder groups rather than to compare the disparate experiences of children vs. young adults. Therefore, representativeness of samples of youth was not essential to the study design.[66]

We recruited a convenience sample of youth and their caregivers living in rural settings in Michigan through collaboration with rural health networks, PCPs, and institutional recruitment resources (e.g., recruitment website). Interested youth and caregivers were referred through their providers or responded to study flyers or online recruitment efforts.

Eligibility criteria were: 9-26-year-olds with a home zip code in rural Michigan, who had visited their PCP at least once in the past 18 months and have not received the HPV vaccination; as some of the participants are minors, so do not make their own vaccination decisions, we also recruited their caregivers. The general term, “youth” refers to children, adolescents, and young adults.[67]

#### Data Collection

We conducted focus groups with multiple stakeholder groups to identify influences on vaccine hesitancy among youth and caregivers in rural Michigan. A focus group guide was developed based on similar studies in other settings. The guides were pilot-tested with rural youth in local primary care practices, and questions were subsequently revised.

Focus groups were organized by participant group: children (ages 9-14; n= 2 focus groups), teens and young adults (ages 15-26; n= 2), and caregivers (n= 2). Two moderators (both family physicians) met with small groups of 4-6 participants using the HIPAA-compliant Zoom video conferencing platform. Focus groups were video recorded and transcribed. One member of the research team (SSG) took extensive notes during the focus group, which were then summarized in memos. Transcriptions were de-identified and checked for accuracy. Transcripts and summary memos were compiled for analysis.

#### Data Analysis

We conducted an inductive, thematic analysis[68] as part of a descriptive, qualitative approach[69]. Three members of the research team trained in qualitative approaches (MD, SC, HK) reviewed each transcript to inductively develop a codebook. At least two people (MD, SC, HK) independently applied descriptive codes from the codebook to all transcripts and memos. We then met in pairs to compare and resolve disagreements; remaining disagreements were discussed in larger team meetings (SSG, MD, SC, HK). Throughout analysis meetings, we identified emergent themes and recorded in analytic memos. Themes were discussed, described in summary memos, and linked to illustrative quotes. Some illustrative quotes were lightly edited for readability.

Descriptive analyses were conducted on the survey data. Cronbach’s alpha and Kendall’s tau were calculated for the scales with several component items. Multiple linear regression analyses were run on the primary dependent variable, the stages of change for inoculating age-eligible patients against the HPV. As the sample was small, only the statistically significant predictor variables were included in the regression analyses; no sociodemographic characteristics were included in this relatively homogeneous sample. All statistical analysis were carried out in SAS 9.4 (SAS Institute Inc., Cary, NC, USA).

After qualitative and quantitative analyses were complete, we integrated the findings through development of a joint display. Joint displays in convergent mixed methods designs are used to compare findings across approaches and develop mixed methods inferences. We created a table that positioned qualitative themes alongside quantitative results to enable comparison and look for discordance and concordance in findings.[54]

The study was approved by the IRBMed of the University of Michigan Medical School.

## Results

Thirty two participants participated across 6 focus groups (2 with children, 2 with young adults, and 2 with parents). Participants included 13 children, 11 young adults, and 8 parents (see Table 1). Most were female and white. Through inductive thematic analysis, five themes were developed and were described in detail alongside illustrative quotes below (see Appendix 1 for additional illustrative quotes).

**Table 1.** Sociodemographics of Focus Group Participants

|  | **N=32** | **Percent** |
| --- | --- | --- |
| **Gender** |  |  |
| Female | 25 | 78. |
| Male | 6 | 19 |
| Non-binary | 1 | 3 |
| **Age** |  |  |
| Child (9-14) | 13 | 41 |
| Young Adult (15-26) | 11 | 34 |
| Parent of participating youth* | 8 | 25 |
| **Race/Ethnicity** |  |  |
| White/Caucasian | 31 | 97 |
| Hispanic/Latino | 1 | 3 |

\*Several parents had more than one child participate in the focus groups

Most rural primary care physicians in the sample practiced in family medicine (71%), followed by internal medicine (12%) and Pediatrics (6%). The primary care physician sample in the survey was two times more female than male (69% vs. 31%, respectively), primarily non-Hispanic White (77%; see Table 2). Most (94%) primary care providers spoke to their patients in English, while one primary care provider communicated in Spanish and English. Most (81%) had attended a US medical school, with the median number of years in practice, 17.5 years. The most common form of practice was either a Physician’s office, single specialty group practice or a Patient Centered Medical Home. The least common practice types were Community Health Centers (Federally Qualified Health Centers, federally funded clinics) and urgent care facilities (one respondent each).

**Table 2.**
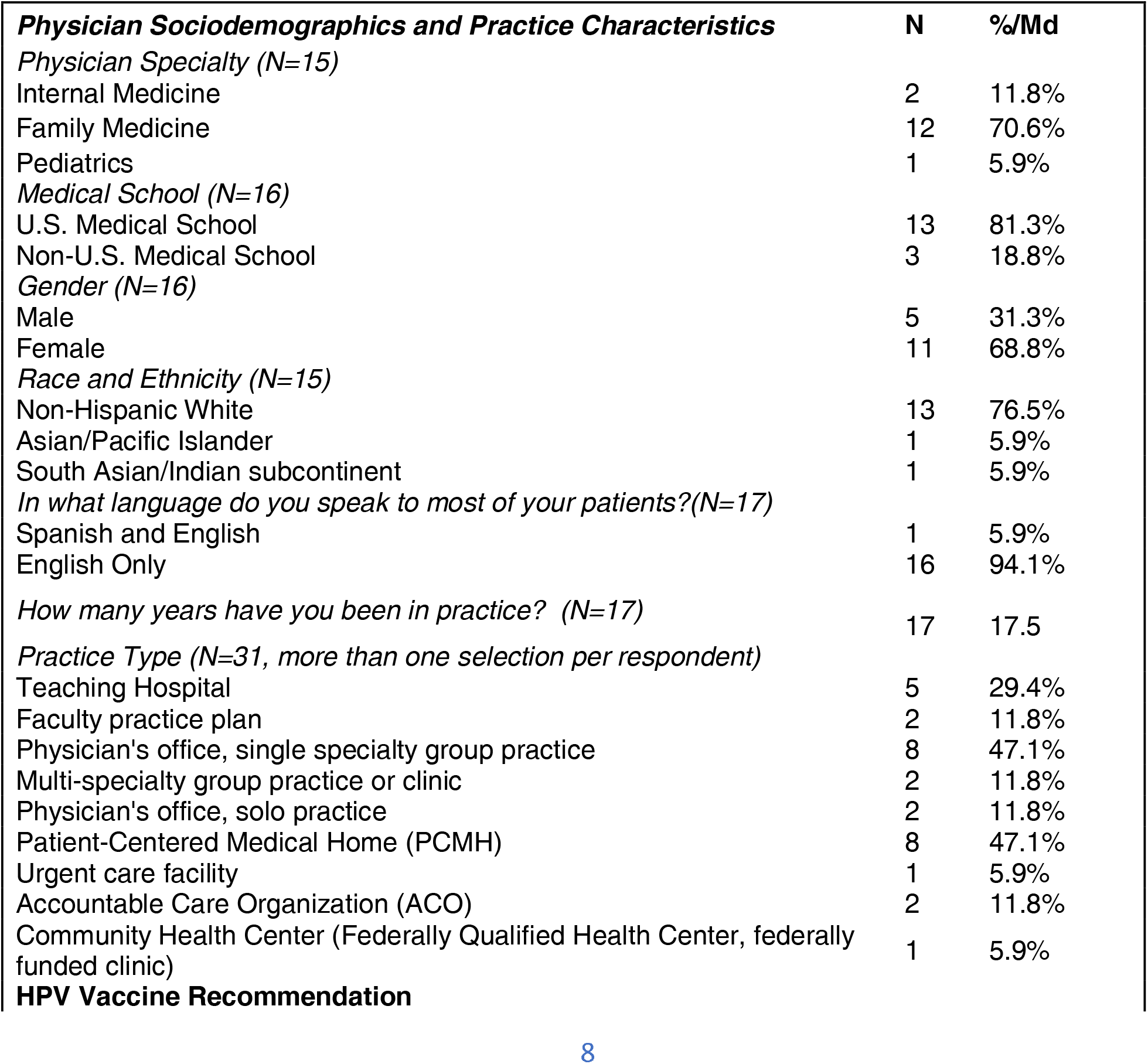
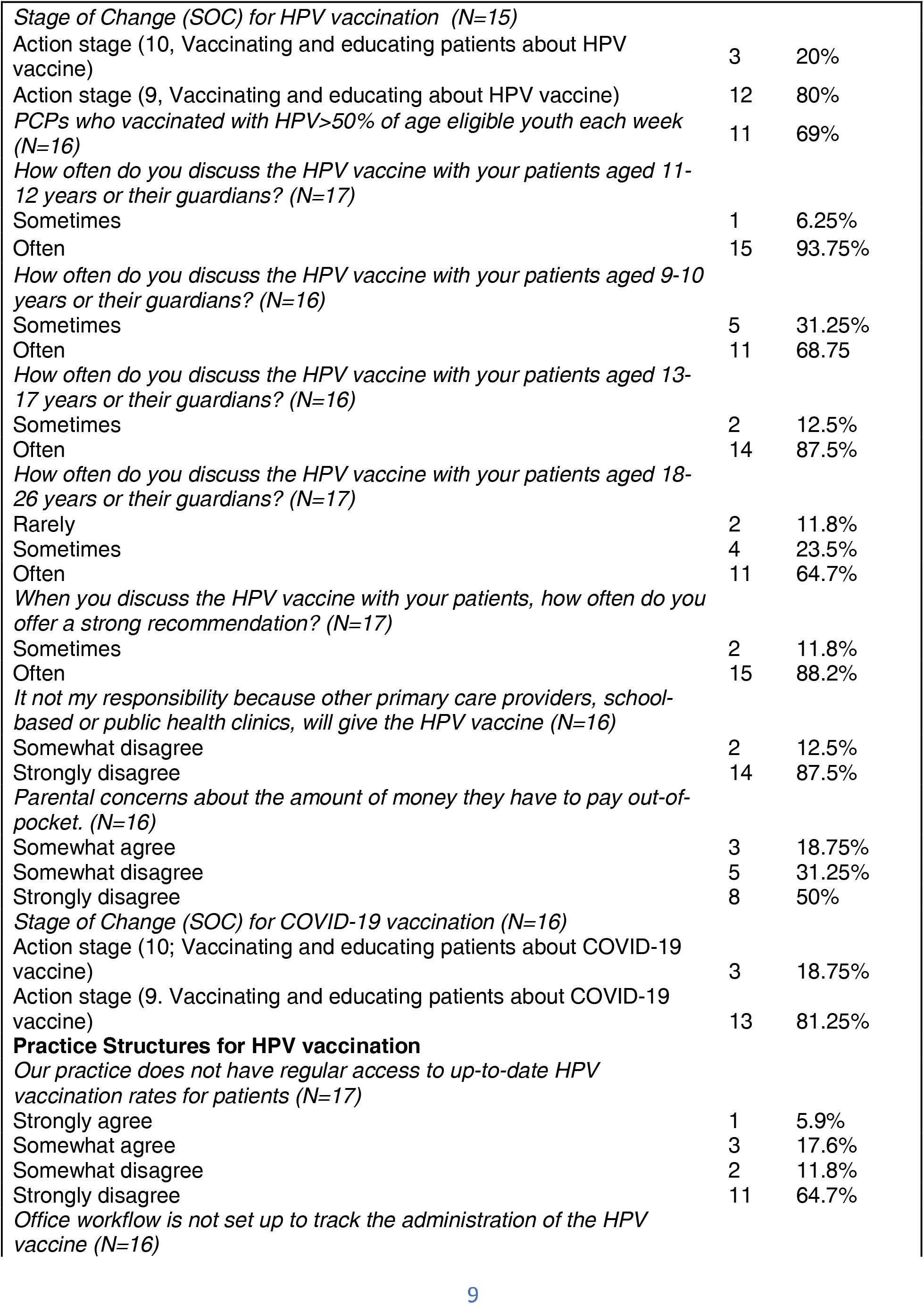
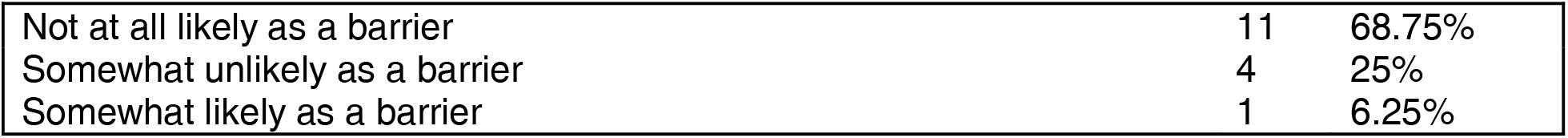
Characteristics of the Primary Care Physician Survey Respondents

Nearly 7 of every 10 respondents respondents reported vaccinating 50% or more of their age-eligible patients each week for HPV. Parental perceived out-of-pocket cost was identified as a barrier to HPV vaccination by 19% of rural PCP’s. All responding physicians report that they are vaccinating and educating their patients about the HPV vaccine. The most frequent discussion about the HPV vaccine was with those age 11-12 or their caregivers/parents (94%) by comparison to those age 13-17 (88%), those 9-10 (69%), or those age 18-26 (65%). The majority (88%) provide a strong HPV vaccination recommendation. The majority (88%) of PCP’s strongly agree that it is their responsibility to vaccinate, rather than the school-based or public health clinics. Regarding clinical structures for vaccination, most (77%) reported having up-to-date HPV vaccination rates. Most (94%) PCP’s reported that office workflows were not a barrier to identify patients for the HPV vaccine. Similar to the HPV vaccine, all respondents reported vaccinating and educating patients about COVID-19 vaccine.

## Theme 1: Limited access and availability of HPV vaccine in rural settings

Overall, both youth and parent participants viewed access and availability to be significant barriers for receiving the HPV vaccine, including access to and availability of the vaccine at convenient locations, insurance coverage and cost, and to a lesser extent, beliefs about healthcare and vaccinations.

Due to their rural setting, transportation created barriers for some participants who would have to drive a fair distance to get the vaccine:

> *“It’s hard for us to get the shots just because where we go for the doctor’s office, is like the 30-minute drive from our house. We don’t have a lot of them like near us”* (Child, Focus Group [FG] 3).

Some who had learned about the HPV vaccine while at their PCP were unable to receive it immediately because it was not stocked. While several youth participants were able to access at their own PCP, they acknowledged that access was a broader barrier for people in their community:

> *“… For us, we can get at a doctor’s office, but I know that certain doctors’ offices don’t necessarily have it. You would have to go to like one of the major hospitals and that can be a challenge for some people”* (Child, FG3).

Though less frequently described in the focus groups, one youth participant highlighted how the transition from pediatric to adult primary care may reduce access for some, as scheduling with a new provider may require a long wait before your first appointment. They described:

> *“I don’t know like [city] is like an hour from me and they’re a lot bigger town, but where I live, now I mean there’s like two doctors’ offices. So, if you’re not already a patient there like half the time it’s like a six-month waitlist so like even if you wanted it you couldn’t just like walk in and get it that week, you would have to wait”* (Young adult, FG2)

Insurance and cost also created barriers for youth and parent participants. First, several young adult participants reported that they did not know whether their insurance included coverage for the HPV vaccination. Second, both youth and parent participants stated that their insurance required receiving the vaccine at somewhere besides the PCP office (e.g., *“I have to go to like a special place to get it, I can’t get [it at] my doctor’s office,”* child, FG3). Participants noted that they were required to get vaccines at the health department, which delayed receipt of the HPV vaccine:

> *“Unfortunately, because of the type of health insurance that we have we can’t get it in his office, if we were cash paying, or if we had a different type of health insurance, we could have just gotten it that day… um so instead we had to go to the health department, so that becomes a little bit of an issue in a rural area, so we didn’t get it right away”* (Parent, FG4)

Finally, participants described how beliefs about healthcare and vaccinations may be a barrier to HPV vaccine uptake in rural communities, as healthcare utilization is low. As one youth participant summarized:

> “*I just know that people around where I live, just didn’t go to the doctor’s office very often… Like if you have a cold, sleep it off. If you broke a bone, go the hospital, reset it, and don’t go [to] your checkups you know.”* (Young adult, FG2)

## Theme 2: Desire for more, unbiased knowledge

Overall, both youth and parent participants emphasized their desire for more knowledge about the purpose, risks, benefits, safety and efficacy to make informed decisions about getting the HPV vaccine. Most youth participants, particularly the youngest cohort, had very limited understanding about the vaccine. For example:

> *“I mean, I haven’t really heard a lot about benefits or downfalls of not getting it or getting it, and so I would say all I really heard is that it’s supposed to help prevent the cancer, but I haven’t heard like how well it prevents it or anything like that, or even likelihood of getting it”* (Child, FG3).

Similarly, a parent described needing more information about HPV as well as the safety and efficacy of the vaccine:

> *“…the safety of it like how many people don’t have reactions. And then um like how effective it is. I don’t even know I guess how prevalent HPV is anymore, like I haven’t researched in many, many years, so I don’t even know like you know, is it worth it, risk with all of that”* (Parent, FG6)

More specifically, both youth and parent participants expressed the need for unbiased and accurate information that emphasizes how this vaccine can prevent HPV and cancer in the future. One parent explained:

> “*For me, I think that if marketing and information and campaigning was from the beginning was more geared towards what it’s actually preventing you know the vaccine is for HPV, but it seems like all of your marketing materials and informational stuff is this is going to help prevent cancer, but it’s not really preventing cancer it’s helping prevent you know infection that could possibly cause cancer down the line. So for me it’s felt like you’re trying to get me to vaccinate for something that is different than what it’s actually intended for if that makes sense”* (Parent, FG4)

Overall, participants expressed a lack of guidance or in-depth conversations surrounding the HPV vaccine from their PCPs. Several youth participants described hearing about the vaccination only briefly (e.g., *“Mostly our doctors said something about it, like saying we should get it”* Child, FG3), but described the interactions as less informative. Often, participants indicated only being given a handout, having brief conversations, or having to advocate for themselves when it came to decision-making for the HPV vaccine. One participant explained:

> *“You know when you’re in the doctor’s office and they kind of spring it on you and you don’t know a lot about it, that’s not the time to make a decision and the next minute and a half, while they’re standing there waiting for you to say yes or no”* (Parent, FG4).

Participants also noted that the presentation of initial information regarding the HPV vaccine and immediate need for a decision in the same appointment made them feel rushed and as though only a weak recommendation was being provided by their healthcare provider.

## Theme 3: Inclusion of youth in decision-making processes

There were mixed reports of whether the communication that did occur with PCPs was directed toward the family and child or only the parent. One parent described how their physician did not include her child, making her question the importance of receiving the HPV vaccine.

> *“It felt like the doctor was talking to almost just me and not including my kid. And I’m somewhat of the mindset, obviously with the exception of when they are newborn babies, with all that stuff that’s their body their choice… but the doctor didn’t want to seem to include them in on that conversation, so that we all kind of left like it, I guess, we don’t need a vaccine for this one”* (Parent, FG4).

Only one parent described a positive interaction with direct and engaging conversation with provider, family, and child.

Participants (both youth and parents) expressed that children, adolescents, and young adults should be included in the decision-making process for the HPV vaccine. Both groups indicated that providers should provide information about the vaccine to both youth and their parents. In addition, if there is a disagreement about the vaccine, most participants indicated that the final decision should be made by the child/young as “it is their body.”

Many youth participants expressed that regardless of their age young people should be allowed to make a decision about the HPV vaccine independently: *“I [think] that young people should be able to make it for themselves, just because it’s our body our choice kind of thing”* (Child, FG3). However, others (both parents and youth participants) expressed that the age of 12-13 when youth typically begin receiving the HPV vaccine, might be too young to be making the decision alone:

> “*So I feel like I feel like to make that own choice you’d have to be a bit older. Because at my age we’re a bit closer to that we’re almost adults anyways so like independent decision making, but like 9-10 11-year-olds, aren’t always the most trustworthy people to make their own decisions like that”* (Child, FG3).

Parents mostly expressed that youth should be included in the final decision on whether they should get the HPV vaccine.

> “*She started getting offered [the HPV vaccine], I think, as a pediatrician around 12 also and then I gave her the choice because it’s not my body, I’m not going to choose that for a teenager*” (Parent, FG6).

Parents also admitted that shared decision making could be more difficult because discussions around the HPV vaccine often include discussions about sexual activity or because parents might feel that an endorsement for the vaccine was also an endorsement to be sexually active.

> “*So I kind of talked about it with my daughter. I explained it was a sexually transmitted disease, so we had you know more talks about what that was. So that that was fun*” (Parent, FG6).

A small number of the youngest cohort also voiced concern about the vaccine related to sexual activity and conversations about sexual activity: “*I also think I can’t talk about it, cause I’m 12. Way too young for that stuff*” (Child, FG5).

## Theme 5: Untapped influence of schools

In general, younger participants agreed that the HPV vaccine has not been openly discussed throughout their education or with their peers (e.g., “*I guess it’s not been a conversation that any of us have brought up”* (Child, FG3)). Most of the youth participants expressed how useful it would have been to learn about HPV during their health education in school. For example:

> *“So I think it could be sorta beneficial like it is in like health classes and that to talk about more variety of different diseases and viruses that people can get like this because I mean where we go to school, we haven’t really talked about much stuff…”* (Child, FG3)

Parents agreed that there is a lack of conversation surrounding the HPV vaccine in schools and within their communities, but there were mixed opinions on whether sexual health should be taught in schools. For example, some of the parents felt that schools may be helpful for unbiased information about sexually transmitted infections and preventive health care because *“sometimes parents don’t know how or where to find information about things*” (Parent, FG4). In addition, schools may be the only place where a child is able to learn and have an open discussion around this topic:

> “*I sometimes have seen families not really being involved in their children’s lives, unfortunately, so short of a quick Google search that might give them loads of misinformation, schools might be the only resource a child has to get that kind of information”* (Parent, FG4).

However, participants did not want schools to push particular messages about the vaccine or require vaccination:

*“But I think it would be cool to see like maybe some kind of education on it, like in like sex ED or something and like a like a take home flyer like, if you want to ask your parents about it kind of thing like partner back, I guess, with the parents, not just saying oh like you have to have it, but start a discussion about”* (Young adult, FG2).

Others felt that schools may not be the appropriate place to talk about sexual health:

> *“I know the whole sexual transmitted disease thing freaks people out a lot of people are waiting for school to tell them about it, which isn’t a good idea in my book”* (Parent, FG6).

Similarly, one parent believed that schools may not be able to effectively educate youth about the HPV vaccine:

> *“I think we almost have to quit relying on the schools to provide information when it comes to STDs and ways of prevention, because, like I said before they don’t really provide enough information, in my opinion, they just gloss over the bare minimum and they’re so scared of scaring the children that or offending anybody that they just gloss over everything um… I’’d rather give an overabundance of information, then have them be uninformed or misinformed”* (Parent, FG6)

## Additional Findings: Comparison to the COVID-19 vaccine

Compared to their experience with the HPV vaccine, participants reported that the COVID-19 vaccine was much more accessible and not subject to the same insurance restrictions, and youth were able to get the COVID-19 vaccine at local pharmacies and pop-up clinics. Still, some parents reported barriers to access for the COVID-19 vaccine in their rural communities, including travel and appointment scheduling.

Though the COVID-19 vaccine was new, participants felt that they had much more information through communication with their providers and public messaging campaigns. For example:

> *“…like with COVID… it has a lot of people talking about it and it like really got around in that, even though it’s [the vaccine] not like out or been known for as, but yet HPV has been around longer, but it seems like no one ever really talks about [HPV vaccine], they just kind of, say, get it or don’t get it and that’s all that is ever said”* (Child, FG3).

Likewise, youth and parents expressed that the COVID vaccine had more immediate, tangible benefits than the HPV vaccine, which made the vaccine an easier decision.

> *“And I think she’s still a little naive to understand all the birds and the bees so. they’re scared of shots so they’re like no I’m not going, and they were like that, with COVID and then as soon as they saw that we couldn’t do anything, when we were in COVID they were like we want a shot and they realized, you know it’s not that bad it was doing it it’s okay move on, so um I feel I have to step in and make that choice for them, because they’re just waiting on a shot and not really all the information that they should know”* (Parent, FG6).

## Integrated Analysis

We juxtaposed the qualitative themes against the findings from the physician survey. The table is a visual joint display, a strategy used for development of integrated findings in a mixed methods study. Joint displays highlight the unique strengths of a mixed methods study by presenting the findings that integrate both quantitative and qualitative results. As described in the revised text, after qualitative and quantitative analyses were complete, we integrated the findings through development of a joint display. Joint displays in convergent mixed methods designs are used to compare findings across approaches and develop mixed methods inferences. We created a table that positioned qualitative themes alongside quantitative results to enable comparison and look for discordance and concordance in findings om Table 3. In Table 3, we describe the survey findings alongside the qualitative themes, using percentages, kendall tau correlation coefficients, and regression analyses among the predictors. The final regression analysis of the outcome of stage of change for HPV vaccination was statistically significant (F=5.3; p=0.02) and included the following predictors: a strong presumptive recommendation (t=0.45; NS), physician anxiety when not certain about a diagnosis (t=-1.02; NS; one item on the physician uncertainty scale [70]), and regular access in the practice to up-to-date HPV vaccination rates for patients, Parental concerns about the amount of money they have to pay out-of-pocket; establishing an office workflow to identify patients for the HPV vaccine (t=-2.17; p=0.0475; t=-3.35; p=0.0048; t=2.31; p=0.0366, respectively), and the approximate percentage of age-eligible patients with insurance coverage for the HPV vaccine (for example, through the Vaccines for Children program; t=0.29; NS). Two clinic structures were associated with increased HPV vaccine uptake (regular access to up-to-date HPV vaccination rate for patients, and office workflows to track HPV vaccine administration).

**Table 3.** Comparison of Qualitative Themes with Quantitative Findings from PCP Surveys

| Qualitative Themes from focus groups with youth and parents/caregivers | Quantitative Findings from PCP surveys |
| --- | --- |
| **Limited access to and availability of HPV vaccine in rural settings**<br>Subtheme:<br>"I also feel like availability could be part of it, because I know, like the only place, I really know you can get it is that, like your doctor's office." (Child, Focus Group 3).<br>Subtheme:<br>"Because they are state funded healthcare they have to go to the health department and and we are in a rural area, so it makes it a lot harder it scheduling more appointments it's taking them out of school or out of extracurriculars or things like that." (Parent, | Parental concerns about the out-of-pocket cost, identified as a barrier to HPV vaccination was a significant predictor of lower HPV vaccine uptake ($t=-3.35$, $p=0.0048$) |
| Focus Group 4).<br>“Unfortunately, because of the type of health insurance that we have we can't get it in his office, if we were cash paying, or if we had a different type of health insurance, we could have just gotten it that day... um so instead we had to go to the health department, so that becomes a little bit of an issue in a rural area, so we didn't get it right away.” (Parent, FG4) |  |
| **Desire for more, unbiased knowledge**<br>““For me, I think that if marketing and information and campaigning was from the beginning was more geared towards what it's actually preventing you know the vaccine is for HPV, but it seems like all of your marketing materials and informational stuff is this is going to help prevent cancer, but it's not really preventing cancer it's helping prevent you know infection that could possibly cause cancer down the line. So for me it's felt like you're trying to get me to vaccinate for something that is different than what it's actually intended for if that makes sense.” (Parent, FG4) | One hundred percent of physicians report that they are vaccinating and educating their patients about the HPV vaccine. These findings suggest a gap between physicians' reported behaviors and patients' need for more, and unbiased, knowledge about the vaccine. |
| **Inclusion of youth in decision-making processes**<br>“...and then I gave her the choice because it's not my body, I'm not going to choose that for a teenager.” (Parent, FG6) | In the regression analysis, a strong recommendation was not associated with HPV vaccination. |
| **Untapped influence of schools**<br>“But I think it would be cool to see like maybe some kind of education on it, like in like sex ED or something and like a like a take home flyer like, if you want to ask your parents about it kind of thing like partner back, I guess, with the parents, not just saying oh like you have to have it, but start a discussion about.” (Young adult, FG2).<br>Subtheme:<br>“So if we're going to educate them about HPV and how it could happen and how to prevent certain things from happening. I feel like the whole dialect of sex education has to change, | The majority (88%) of PCP's strongly agree that it is their responsibility to vaccinate, rather than the school-based or public health clinics to administer the HPV vaccine. |
| where it's not abstinence only; that's very one track minded." |  |
| **Clinical structures to track and workflows to vaccinate age-appropriate youth**<br>Subtheme:<br>"...And you knew this appointment was coming up why doesn't why don't you have it [the HPV vaccine]..." (Parent, FG4). | Clinics with up-to-date HPV vaccination rates for patients ($t=-2.17$, $p=0.0475$) were significantly associated with increased HPV vaccine uptake. Those offices with established workflows to identify patients for the HPV vaccine were significantly associated with higher HPV vaccine uptake ($t=2.31$; $p=0.0366$). |
| **Comparison to the COVID-19 vaccine**<br>"...but yet HPV has been around longer, but it seems like no one ever really talks about [HPV vaccine], they just kind of, say, get it or don't get it and that's all that is ever said." (Child, FG3). | Stage of change to administer the covid-19 vaccine was not associated with stage of change to administer the HPV vaccine, however ($t=0.59$; NS). |

## Discussion

Interestingly, the qualitative and quantitative data reveal two different perspectives on the role of the physician in educating about vaccination, on involvement of youth in decision-making about the vaccine, and in the barriers to vaccination. All of the surveyed physicians reported educating their patients about the vaccine, while the focus group participants reported little unbiased information about the HPV vaccine. Physicians identified some internal structural barriers to vaccination, in access to up-to-date lists in the practice, to identify those eligible for a vaccination. Further, primary care physicians reported that vaccination against HPV was their role, not theirs of the school or the public health clinic. Yet, only one focus group participant suggested a community liaison to link the primary care clinic and the unvaccinated community. Both the focus group participants and the primary care physicians, however, perceived out-of-pocket cost or insurance coverage as a barrier to uptake among patients. Cost is seen as a strong barrier despite the wide availability of free HPV vaccines under the Vaccines for Children Program.

While youth in the study sought more involvement in decision-making around the vaccine administration, physicians were most often offering a presumptive announcement, stating that the vaccination would be given at that visit. Strong evidence from a randomized clinical trial on presumptive announcement vs. conversational approaches support the influence of the former on increased HPV vaccine uptake across rural and urban settings [46]. Adolescents and parents in our focus group expressed unease with this method, wanting more information and more involvement in decision making. Interestingly, Beavis, et al.[71], interviewed vaccine hesitant parents of adolescents and found a similar desire for more information, and dissatisfaction with provider encounters, as well as a lack of confidence in their vaccine decision-making.

Further, while older youth (age 18-26) are less likely to receive a recommendation for the HPV vaccine relative to younger youth (age 11-12), the focus group findings revealed their interest in being vaccinated now that they can make those decisions themselves. The youngest participants in the focus groups also expressed an interest in the HPV vaccination decision, as well, however, as did some of their parents. Individuals in rural settings may place a higher value on bodily autonomy than comparable others [37]. Further, our findings support the importance of “familial decision making,” including parents and adolescents/children, when deciding whether or not to vaccinate against HPV [72].

Parental participation in our focus groups suggested that parents felt rushed to decide whether or not to vaccinate at the current visit. Further, youth voiced concerns regarding lack of knowledge and reliable information being given prior to having to make a decision about vaccinating their children for HPV. Lower levels of parental knowledge regarding the HPV vaccine and perceived vaccine efficacy are negatively associated with vaccine initiation [37].

Participants in our focus groups suggested that providers communicate with parents about the upcoming vaccine recommendation and provide education about the vaccine the year prior to the child becoming eligible. A recent survey of urban vaccine hesitant parents[71] also found that introducing the HPV vaccine prior to the appointment, including a list of questions to help parents and youth prepare for primary care visits, in-depth discussions, and written materials could increase both parental and youth knowledge of the HPV vaccine, thus facilitating the decision. To desexualize the vaccine, that is, detach it from decisions about engaging in sexual activity that often arise in adolescence, introducing the vaccine when children are age 9 has been found through preliminary studies to increase uptake, and seems well-accepted by parents [73–75].

The mixed method approach combines the strengths of both quantitative and qualitative research, by allowing the researcher to add insights that could be omitted when only a single method is adopted. The rigorous integration of the qualitative and quantitative analyses is a major strength of the study research questions, the method, analyses, and the findings. The study does have limitations, however. All focus groups were remote, so participation was limited by connection capability and speed. Remote data collection may increase participation among those who may otherwise face logistical or time constraints, and is becoming widely accepted for telemedicine. [76, 77] Use of a convenience sample from a self-selected set of focus group participants, and the low rate of rural primary care physician response, may bias the findings and limit generalizability. A two-page consent form required by the IRB discouraged respondents, further reducing the sample, however. While we recruited licensed primary care providers in the state, we were unable to verify the sample frame for rural providers. The response rate is comparable to that from another recent physician survey of HPV vaccination practices, however. [78] Participating caregivers were few in number; the findings were consistent across respondents, however, and the constructs were saturated through these responses. As is normative across mixed methods studies, research questions, rather than testable hypotheses, founded the study.[54]

The findings suggest several strategies to decrease rural HPV vaccine hesitancy. Within clinics, integrating HPV vaccine surveillance data with the electronic health record could facilitate more rapid retrieval of vaccination status. The State of Michigan maintains a robust system of monitoring HPV vaccinations, but the data are not necessarily within clinical electronic health records, or on dashboards, so must be retrieved by request. Once available, routine audit and feedback of these data could increase their use by clinical teams. Further, within clinical settings, additional workflow mapping for the HPV vaccine could facilitate routine vaccination.

Among primary care physicians, training in announcing the HPV vaccine, thus integrating it with the other adolescent vaccines, could decrease hesitancy [79]. Practice facilitation and academic detailing with clinicians, front office staff, and others could facilitate redesign for HPV vaccination [80–83]. This redesign could include identifying community-based resources like community pharmacies that are enrolled in the Vaccines for Children program [48, 84], that could increase access in rural areas. Additional education of parents and youth prior to the visit could facilitate confidence in their vaccine decision-making. Developing local champions among both primary care providers and rural youth for the HPV vaccine could enhance decision-making skills, disseminate accurate and unbiased information, and increase trust in the vaccine [80].

Further study to examine the gap between provider vaccination recommendations, decision-making factors for parents and youth, and vaccination outcomes is warranted. The influence of community and policy factors on this decision-making is a next step.

## Author Contributions

Conceptualization: SSG. Methodology: SSG, MD, CJ, AK. Formal Analysis: SSG, MD, DC, SC, HK. Resources: SSG, AK, MS, CJ, AK. Data Curation: Writing— Original Draft, Preparation: SSG, MDJ, SC, HK, DC. Writing—Review and Editing: SSG, MDJ, CJ, AK, MS, SC, HK, RH. Supervision: SSG, MDJ. Project Administration: SSG. Funding Acquisition: SSG.

## Institutional Review Board Statement

The study was conducted according to the guidelines of the Declaration of Helsinki, and approved by the Institutional Review Board of the University of Michigan Medical School (IRBMed) (protocol HUM00183900, approved 8/22/2023), and the MyMichiganHealth, University of Michigan Health Systems IRB (formerly, Mid Michigan Health).

## Informed Consent Statement

Written informed consent was obtained from all participants involved in the study.

## Data Availability

Anonymized data produced in the present study are available upon reasonable request to the authors

## Acknowledgements

An earlier version of this paper was presented at the 2023 NAPCRG Annual Meeting. We thank colleague Dr Diane Harper, University of Michigan School of Medicine for presenting the paper at the 2023 NAPCRG Annual Meeting. We appreciate the recruiting support of the following MSU/MyMichigan Medical Center physicians: Drs. Michelle Nelson (attending and supervisor), Jonathan Erius, Daniel Gross, and Daniel Kim. We appreciate the assistance of Meghann Lewis with the recruitment of participants and scheduling of the focus groups. We thank Yuhong Zhang, research data manager, and Courtney Olson, for preparation of the tables.

## Appendix

### Appendix 1. Themes, sub-themes, and additional illustrative quotes

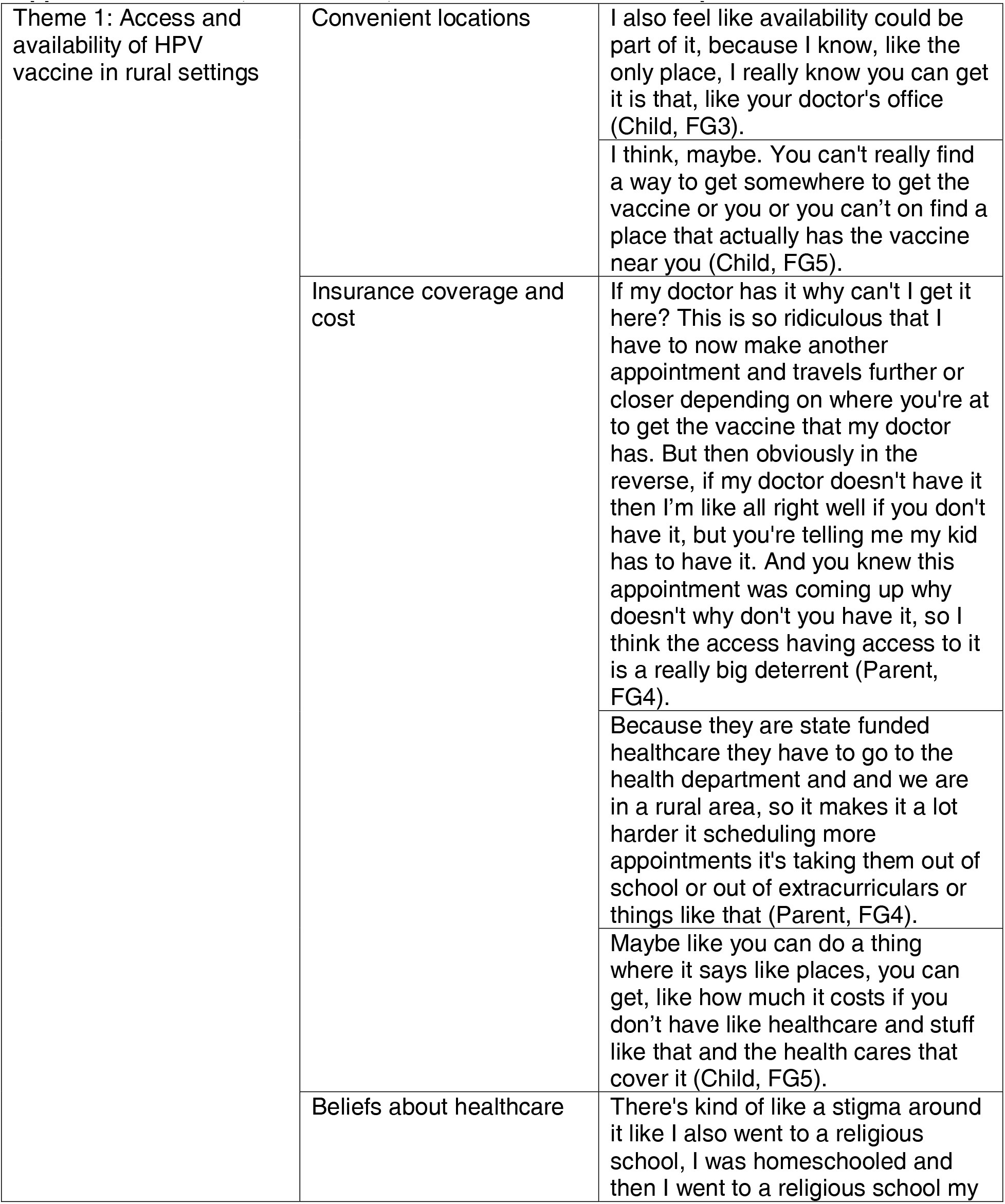
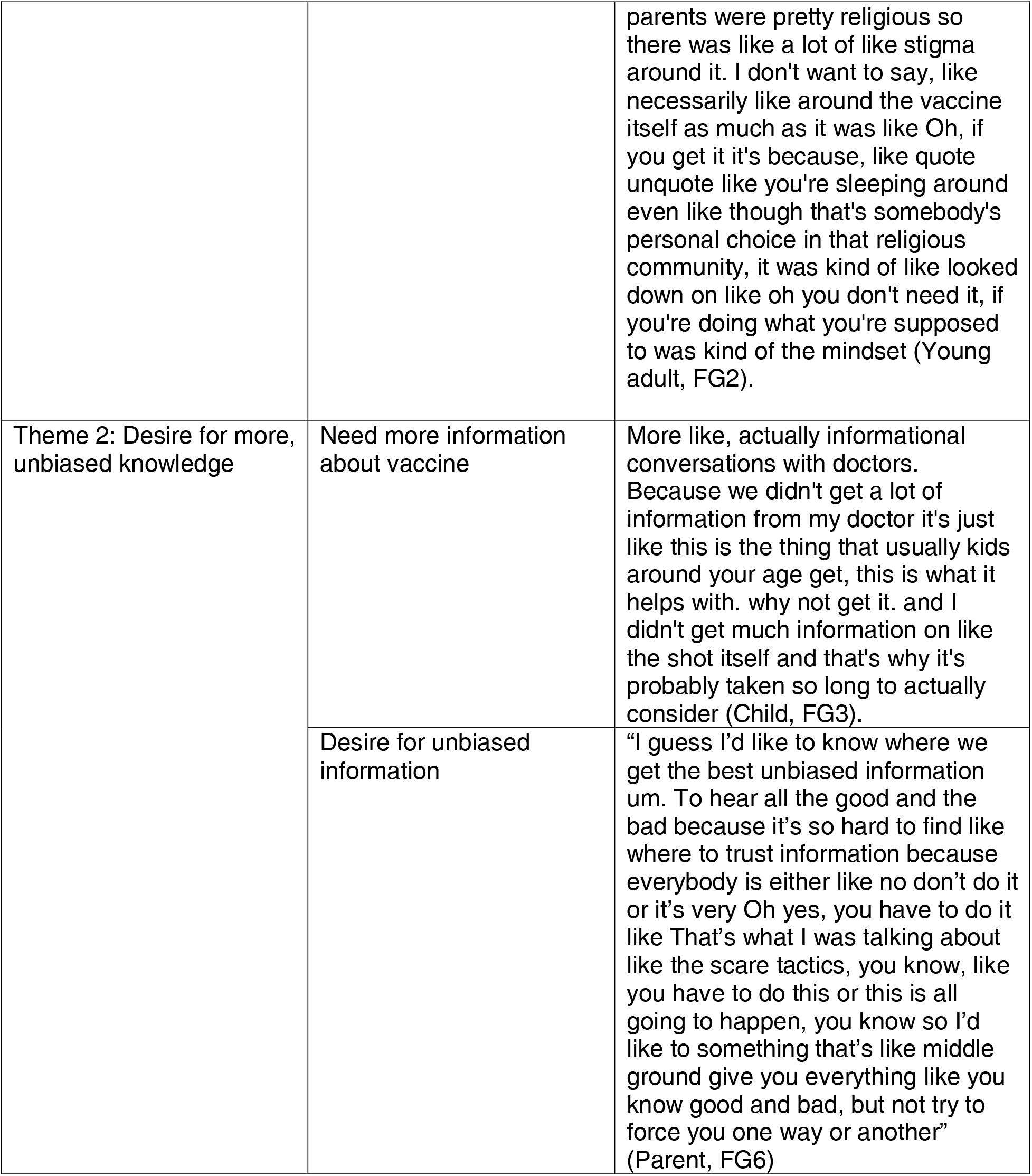
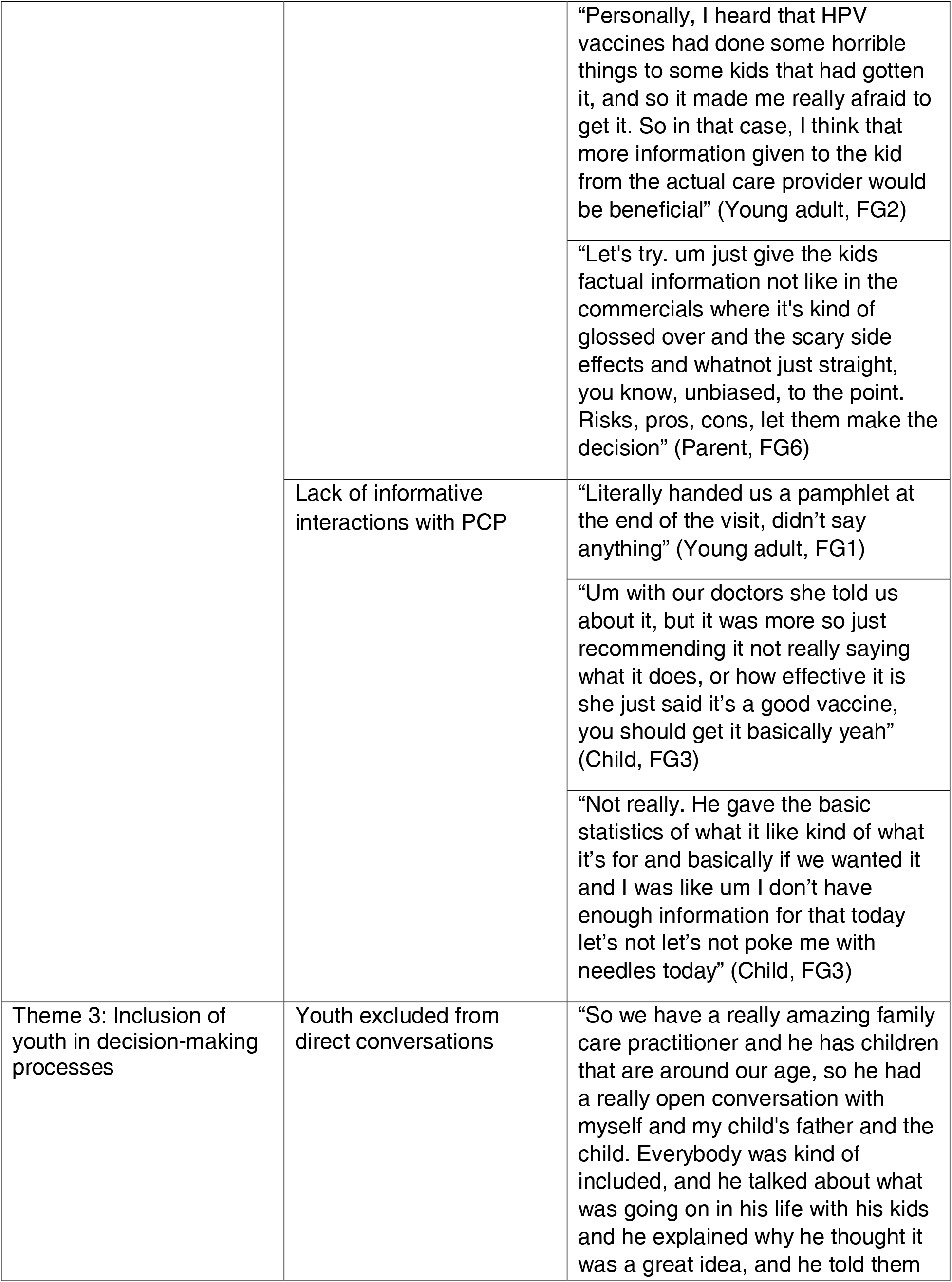
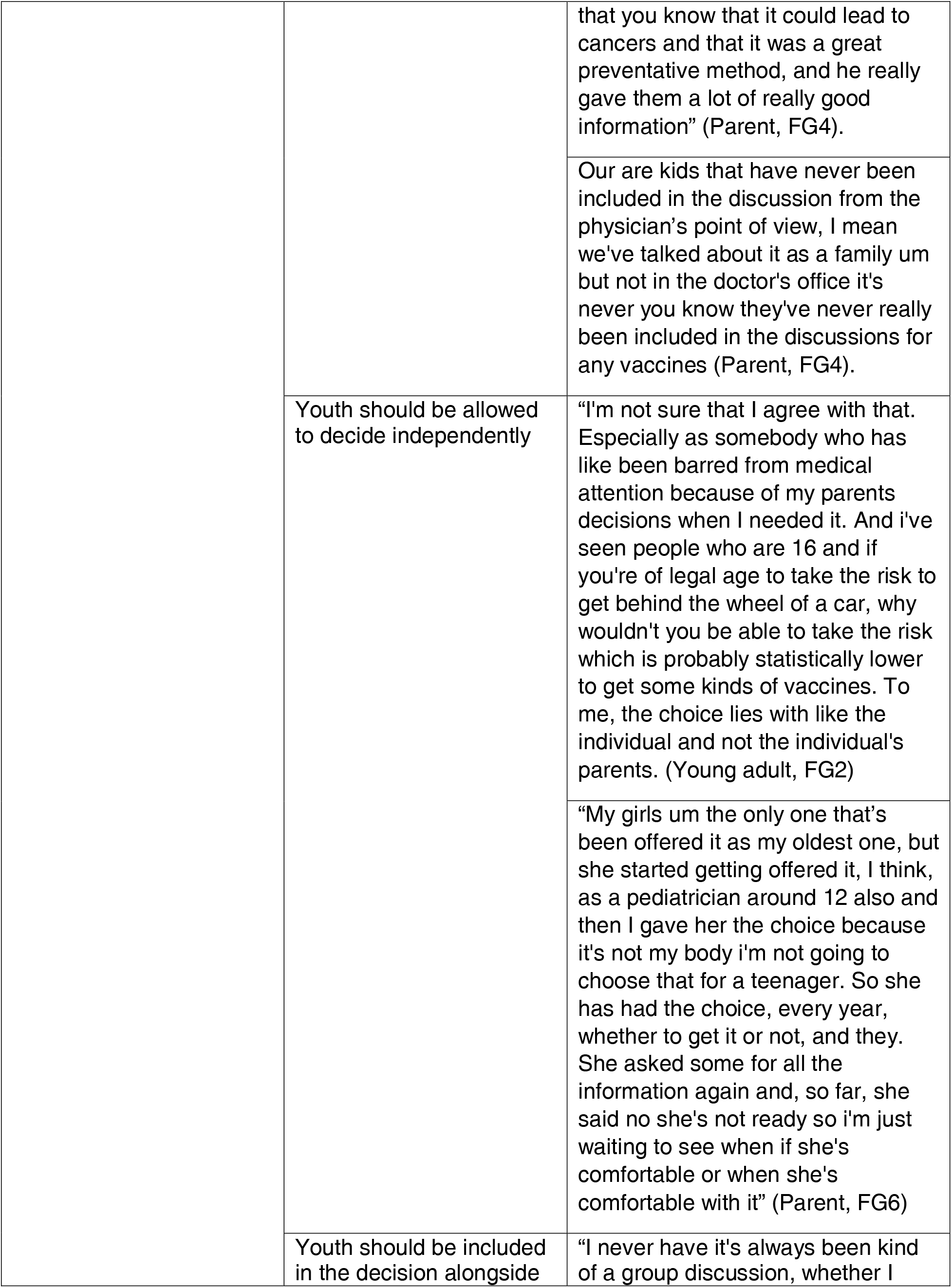
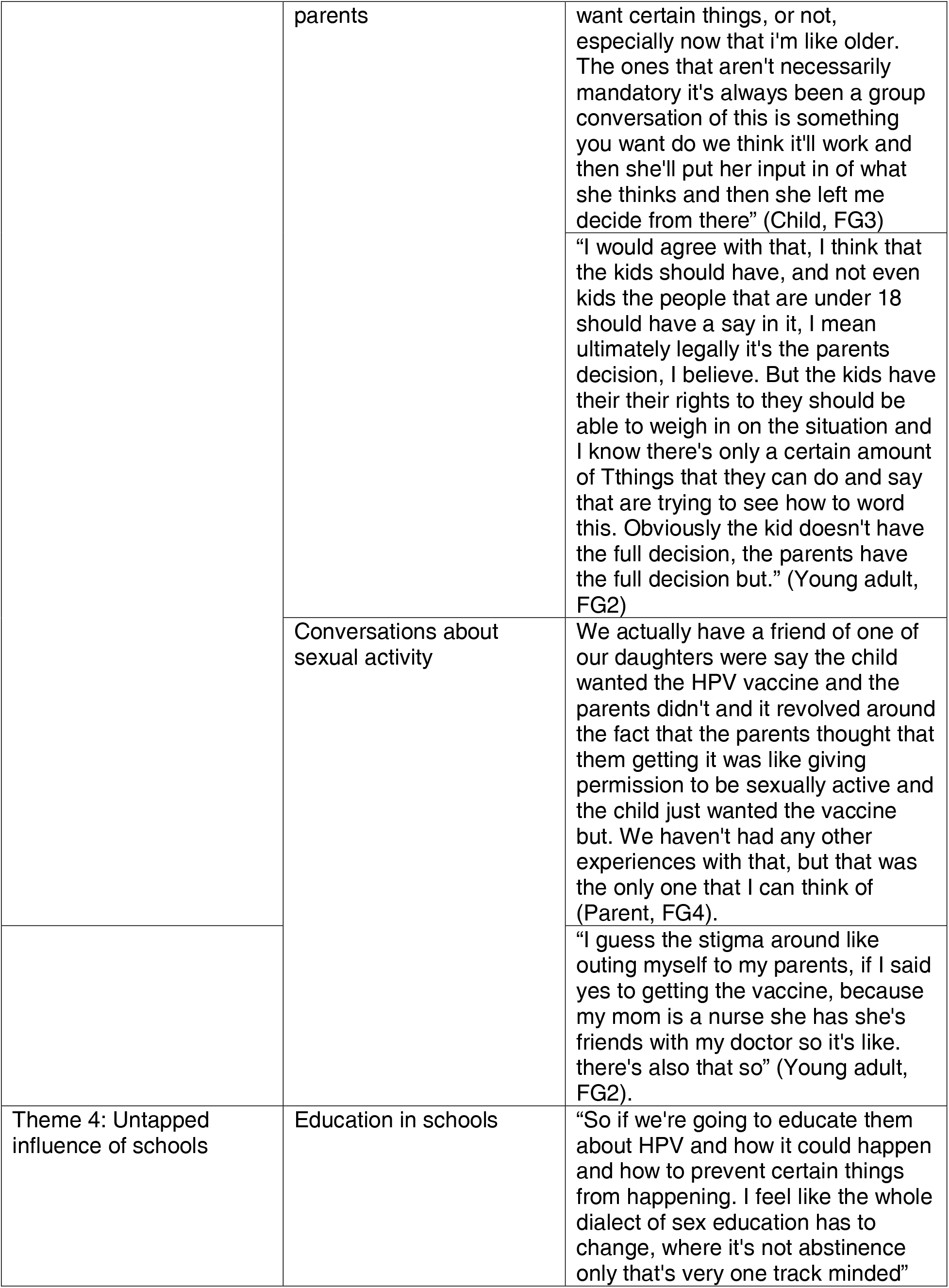
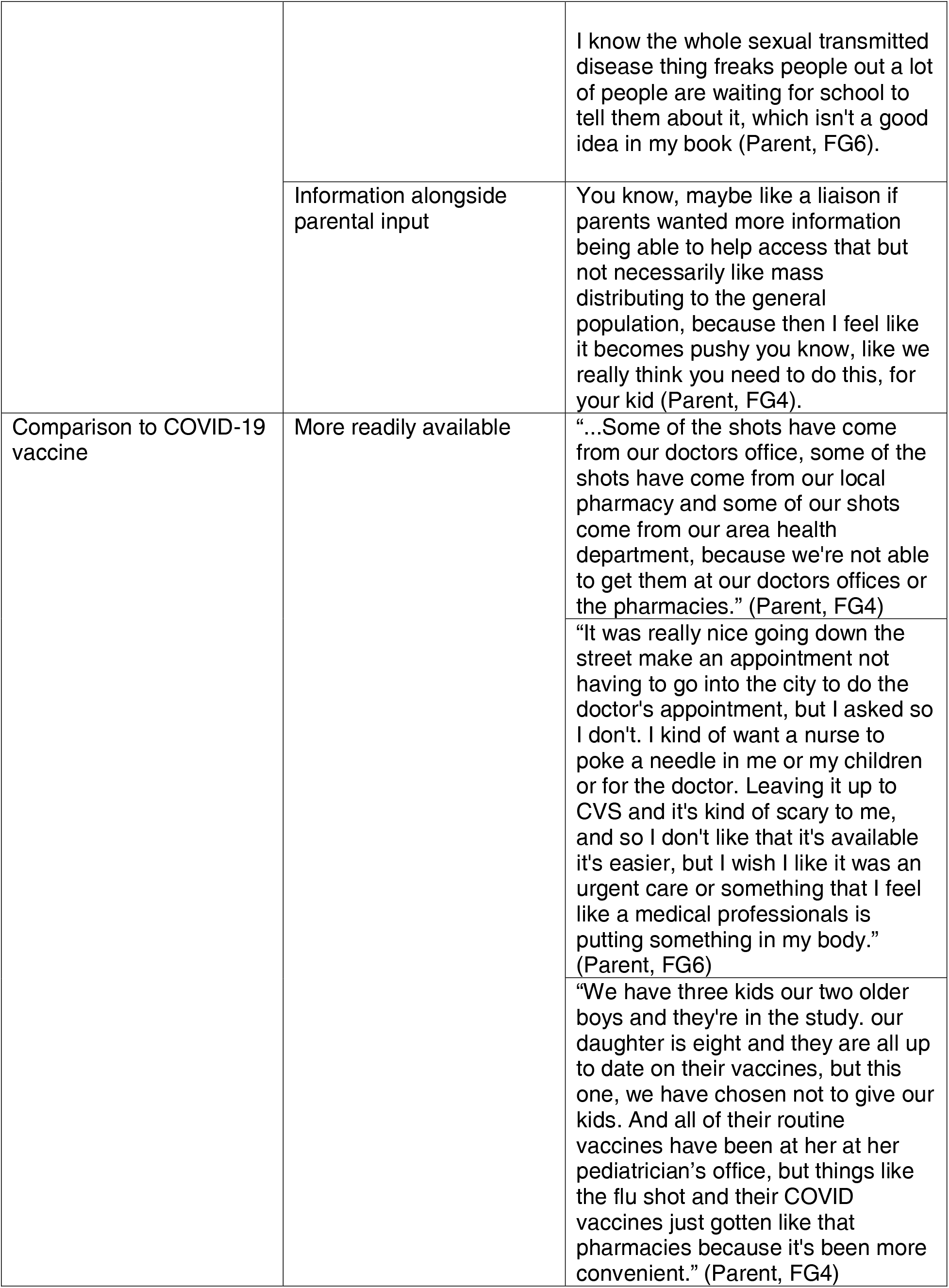
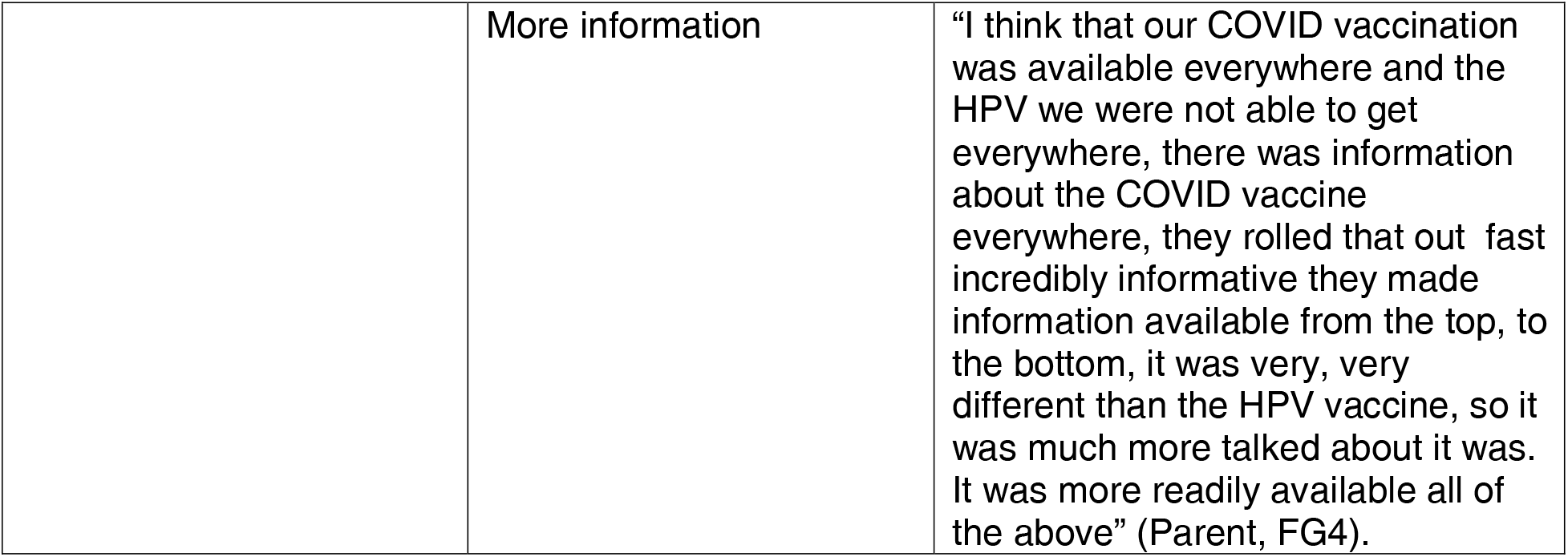

